# Matching Heterogeneous Cohorts by Projected Principal Components Reveals Two Novel Alzheimer’s Disease-Associated Genes in the Hispanic Population

**DOI:** 10.1101/2025.01.18.25320774

**Authors:** Julian Daniel Sunday Willett, Mohamad Waqas, Serhiy Naumenko, Kristina Mullin, Julian Hecker, Lars Bertram, Christoph Lange, Ioannis Vlachos, Winston Hide, Rudolph E. Tanzi, Dmitry Prokopenko

## Abstract

Alzheimer’s disease (AD) is the most common form of dementia in elderly, affecting 6.9 million individuals in the United States. Some studies have suggested the prevalence of AD is greater in individuals who self-identify as Hispanic. Focused results are relevant for personalized and equitable clinical interventions. Ethnicity as a stratifying tool in genetic studies is often accompanied by genomic inflation due to heterogeneity. In this study, we report GWAS and meta-analyses conducted among NIAGADS subjects who self-identified as Hispanic and All of Us (AoU) sub-cohorts matched to that cohort, using projected genetically-derived principal components, with and without age and sex. In Hispanic NIAGADS subjects, we identified a common variant in *PIEZO2* that was protective for AD with a p-value just beyond genome-wide significance (p = 5.4 * 10^-8^). Meta-analyses with genetically-matched AoU participants yielded three (two novel) genome-wide significant AD-associated loci based on rare lead variants: rs374043832 (*RGS6/PSEN1*), rs192423465 (*ASPSCR1*), and rs935208076 (*GDAP2*), which were also nominally significant in AoU sub-cohorts. We thus demonstrate an efficient way to select subjects from large heterogeneous biobank cohorts who are genetically similar to a smaller disease-specific cohort, yielding novel disease-relevant findings.

## Introduction

Alzheimer’s disease (AD) is a high-morbidity illness predicted to affect 13.8 million individuals in the United States by 2050^1^. While studies have revealed meaningful disease-relevant genes^2,3^, fewer studies have clarified its genetic determinants in groups beyond those of European ancestry. AD has been observed to be more prevalent in Latinos with several genes linked to familial AD risk in this group, including *PSEN1* and *BIN1*^4^. Polygenic risk score calculations for Hispanic participants have found APOE alleles to be useful in predicting mild cognitive impairment^5^. A GWAS was conducted on the Hispanic Community Health Study in 2021, identifying a novel variant with association with mild cognitive impairment^6^.

Ethnicity is an imperfect stratifier in genetics studies due to the additional diversity it can fail to capture. Beyond the National Academies releasing a report in 2023 acknowledging the limitations of social-constructed subgroup analyses in genetics^7^, individuals who identify as Hispanic are incredibly diverse with significant genetic variation across communities^8^, contributed to by historical migration and admixture^9,10^. This heterogeneity, which can produce genomic inflation and reduce confidence in results, is not unexpected considering that ethnicity captures fewer generations of one’s family history than genetic ancestry, which groups individuals by the predicted geographic distribution of one’s ancestors from millennia before. Ethnicity as a stratifier remains common in AD genetics studies^11,12^, perhaps driven by efforts to yield results that can generalize to individuals of diverse backgrounds. This complicates efforts to combine different studies of Hispanic subjects together in a meta-analysis, given possible heterogeneity.

In this study, we aimed to develop a means of studying AD across populations that are historically heterogeneous. We performed a genome-wide association study (GWAS) in a Latino population from NIAGADS with clinical AD and sought to replicate the findings in a large population-based cohort from All of Us (AoU) with an AD-by-proxy phenotype. Combining concepts underlying population genetics and clinical trials, we matched NIAGADS participants who self-identified as Hispanic to participants in AoU using genetically derived principal components, after projecting subjects in AoU onto our NIAGADS sub-cohort PC space. We then ran GWAS on each subgroup and meta-analyzed the results, identifying two novel rare loci associated with AD nominally significant in both sub-cohorts, alongside *APOE* (**Figure 1**). We identified two novel rare AD-associated loci and show how population stratification can be controlled in studies with diverse participants while maximizing power and yielding more generalizable results. We also demonstrate an efficient way to select subjects from a large biobank cohort who are genetically similar to a disease-based cohort.

**Figure 1.**
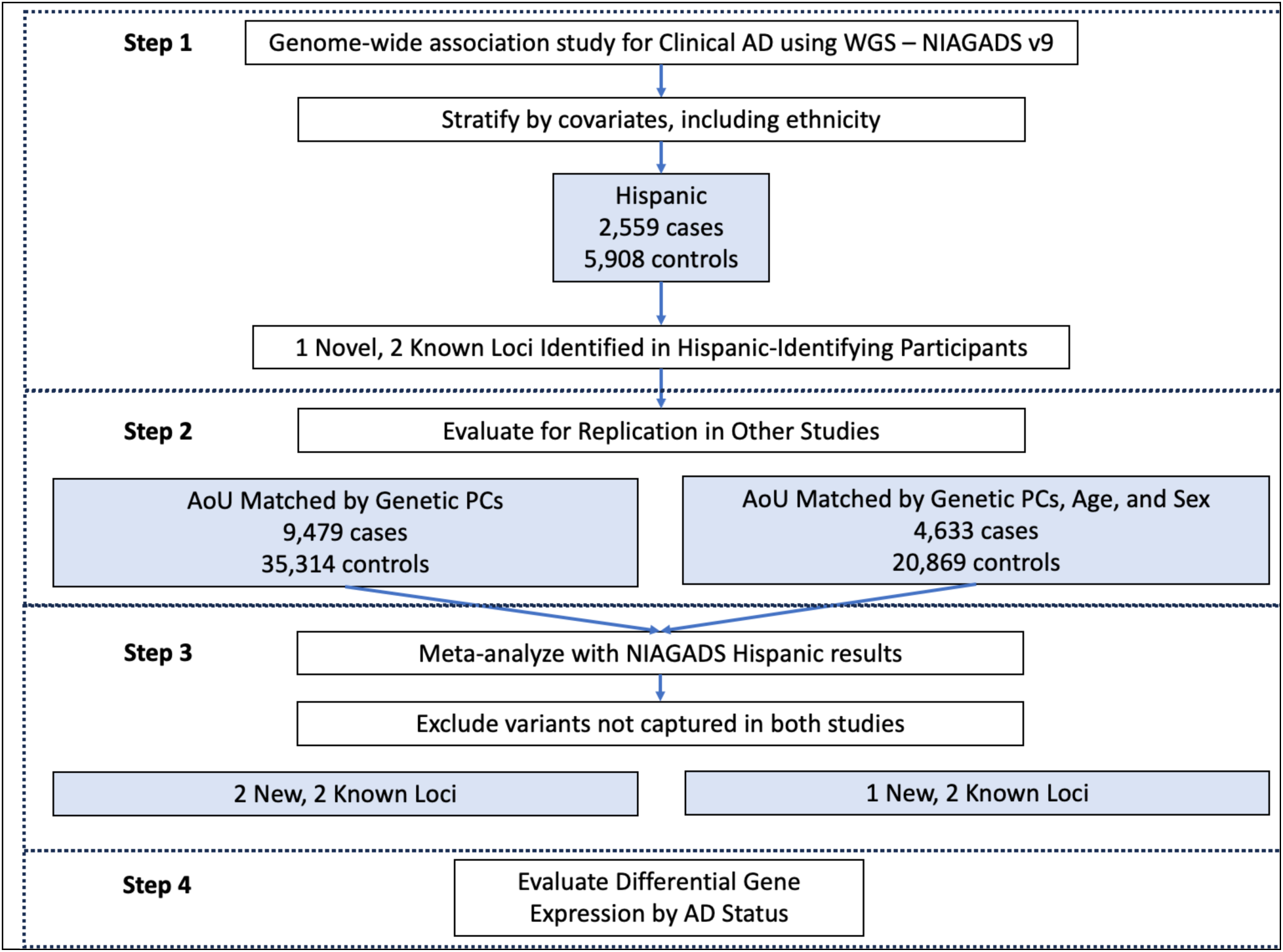
Study design. NIAGADS result replication in All of Us (AoU) was accomplished by matching AoU participants to NIAGADS using solely genetically-derived projected principal components (PCs) or PCs, age, and sex.

## Results

### Sample, Variant-Level Quality Control and Genetic Matching

For NIAGADS, we isolated genetic data for a cohort of individuals self-reporting Hispanic ethnicity. We removed technical replicates, individuals missing an AD diagnosis, related individuals, samples with a high missingness rate, outliers based on principal components, and variants that were outliers based on HET/HOM ratio. This resulted in 38,989,734 variants and 8,467 samples.

Next, we calculated “reference” PCs on our NIAGADS sub-cohort, using a set of LD-pruned common variants present in NIAGADS and All of Us (AoU). We projected AoU PCs onto the reference PCs and used both PC sets to select subjects from AoU which genetically matched our discovery dataset. We created two matching cohorts using subclassification models: the first, matched solely on PCs and Affection Status, the second, also accounting for age and sex. The genetic data for each AoU sub-cohort was then filtered for flagged samples or variants and variants with excessive missingness. This left 49,511,921 variants and 44,793 samples in the AoU sub-cohort, matched by solely projected PCs and 40,635,522 variants and 25,502 samples in the AoU sub-cohort, matched by projected PCs, age, and sex (**Supplemental Table 1**).

For the meta-analysis, matching AoU participants to NIAGADS participants who self-identified as Hispanic increased the case count five-fold when matching with genetic PCs, and almost three-fold when matching with genetic PCs, age, and sex (**Supplemental Table 1**). Including age and sex as covariates yielded an AoU matched participant age distribution more closely resembling NIAGADS with minimal effect on the proportion reporting female sex (**Figure 2**). The distribution of genetic ancestries varied dramatically between NIAGADS participants who self-identified as Hispanic and the matched AoU participants (**Supplemental Table 1 and Supplemental Figure 2**).

**Figure 2.**
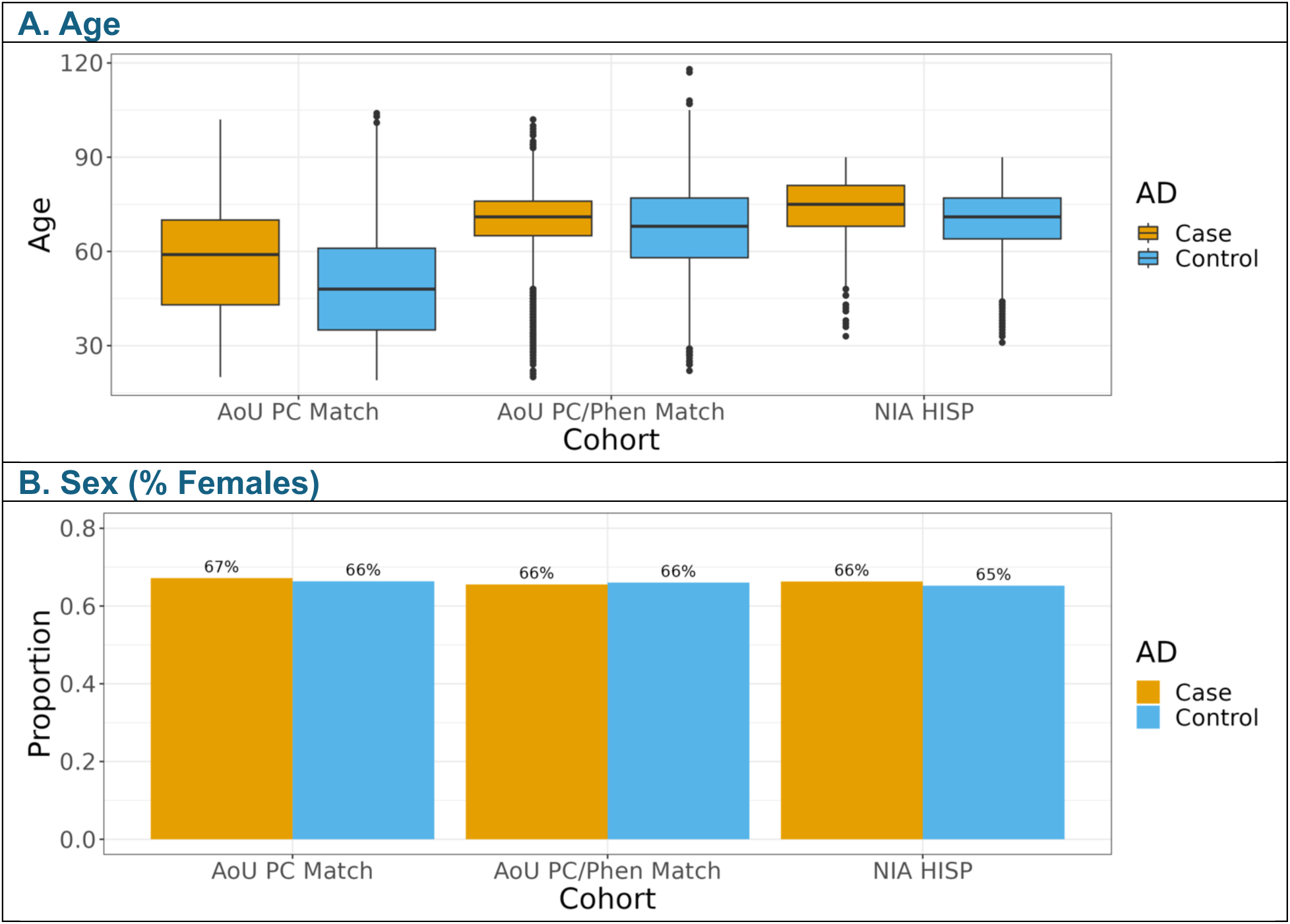
Cohort demographics for All of Us (AoU) and NIAGADS (NIA) studies. AoU PC Match included AoU participants matched to NIAGADS participants who self-identified as Hispanic (NIA HISP) using the first twenty PCs, obtained by projecting AoU array data onto NIAGADS genetic data. AoU PC/Phen Match included AoU participants matched to NIAGADS participants using the first twenty projected PCs, age, and sex. Matching was accomplished using MatchIt’s subclassification method, isolating participants with a minimum weight of 1, matching or exceeding the NIAGADS reference population weight.

Individuals who self-identified as Hispanic in NIAGADS were predominantly predicted to have European genetic ancestry with AoU predominantly composed of participants predicted to have admixed American genetic ancestry (**Supplemental Table 1**). Most AD-by-proxy cases in the AoU sub-cohorts were predicted to have European genetic ancestry, similarly to NIAGADS (**Supplemental Table 1**).

### NIAGADS Genome-Wide Association Study

We identified 3 loci with variants beyond or close to genome-wide significance for NIAGADS participants who self-identified as Hispanic, with the study having minimal evidence of genomic inflation (λ = 1.00) (**Figure 3A, Supplemental Figure 1A**). The first locus was at *APOE*, with the lead variant p-value of 3.2 * 10^-46^. The second locus within the *ZFYVE1* gene, encoding Zinc Finger FYVE Domain-containing Protein 1, resides 138 kb from the early-onset familial AD gene, presenilin 1 (*PSEN1*), and the lead variant had a genome-wide significant p-value of 6.7 * 10^-10^ (**Table 1**). The *ZFYVE1* locus most likely represents the *PSEN1* haplotype from Caribbean Hispanics^13^, observed in a prior NIAGADS release^14^. The third novel locus was at the gene encoding Piezo-Type Mechanosensitive Ion Channel Component 2 (*PIEZO2*), and the lead common variant was protective against AD with a p-value just beyond genome-wide significance (p = 5.4 * 10^-8^) (**Table 1**). *PIEZO2* encodes the pore-forming subunit of the mechanosensitive cation Piezo channel, which is required for mechanically activated currents with roles in sensing touch and tactile pain^15^. *PIEZO2* has homology to *PIEZO1*^16^, which has been previously linked to amyloid beta clearance^17^.

**Figure 3.**
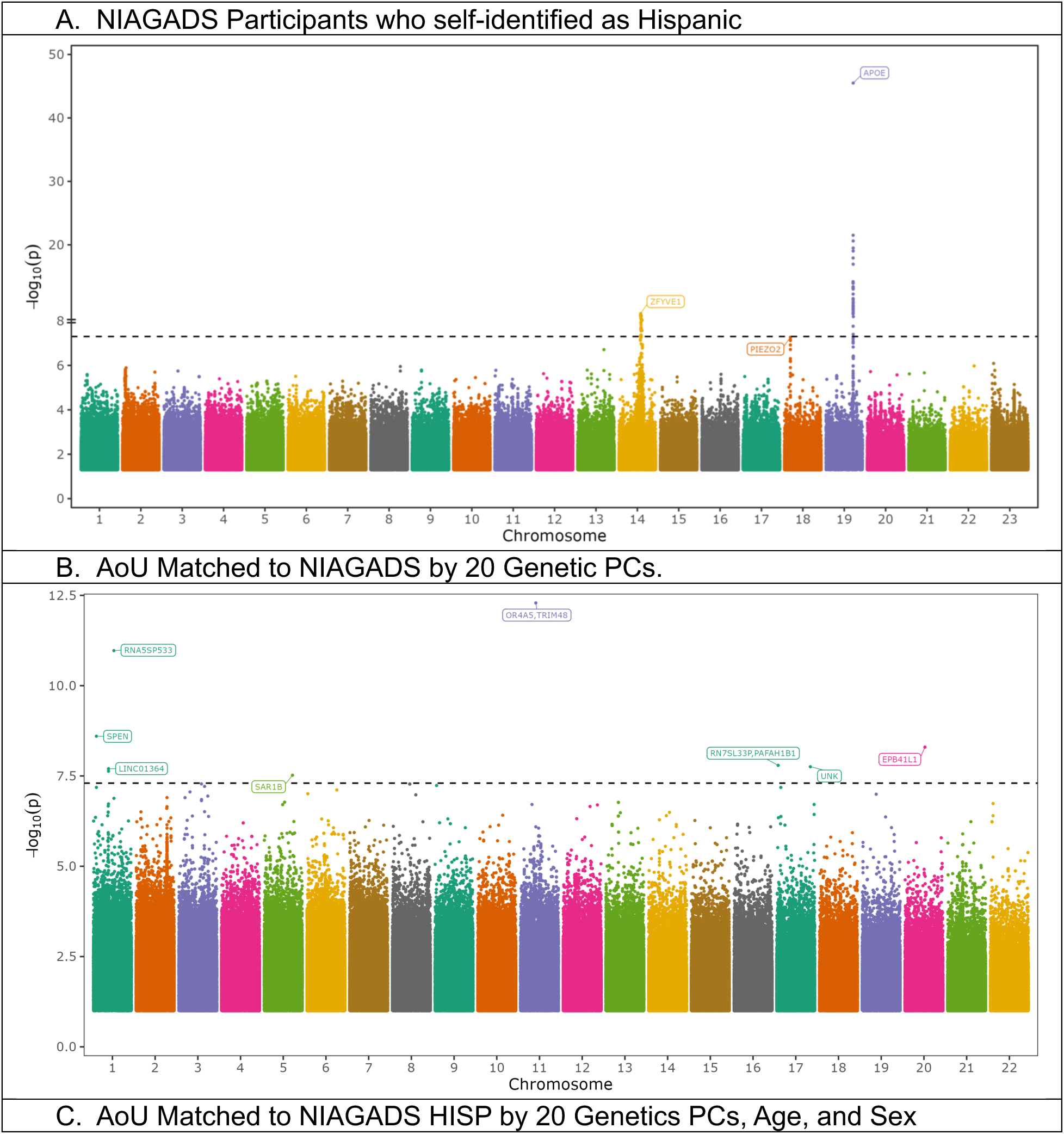
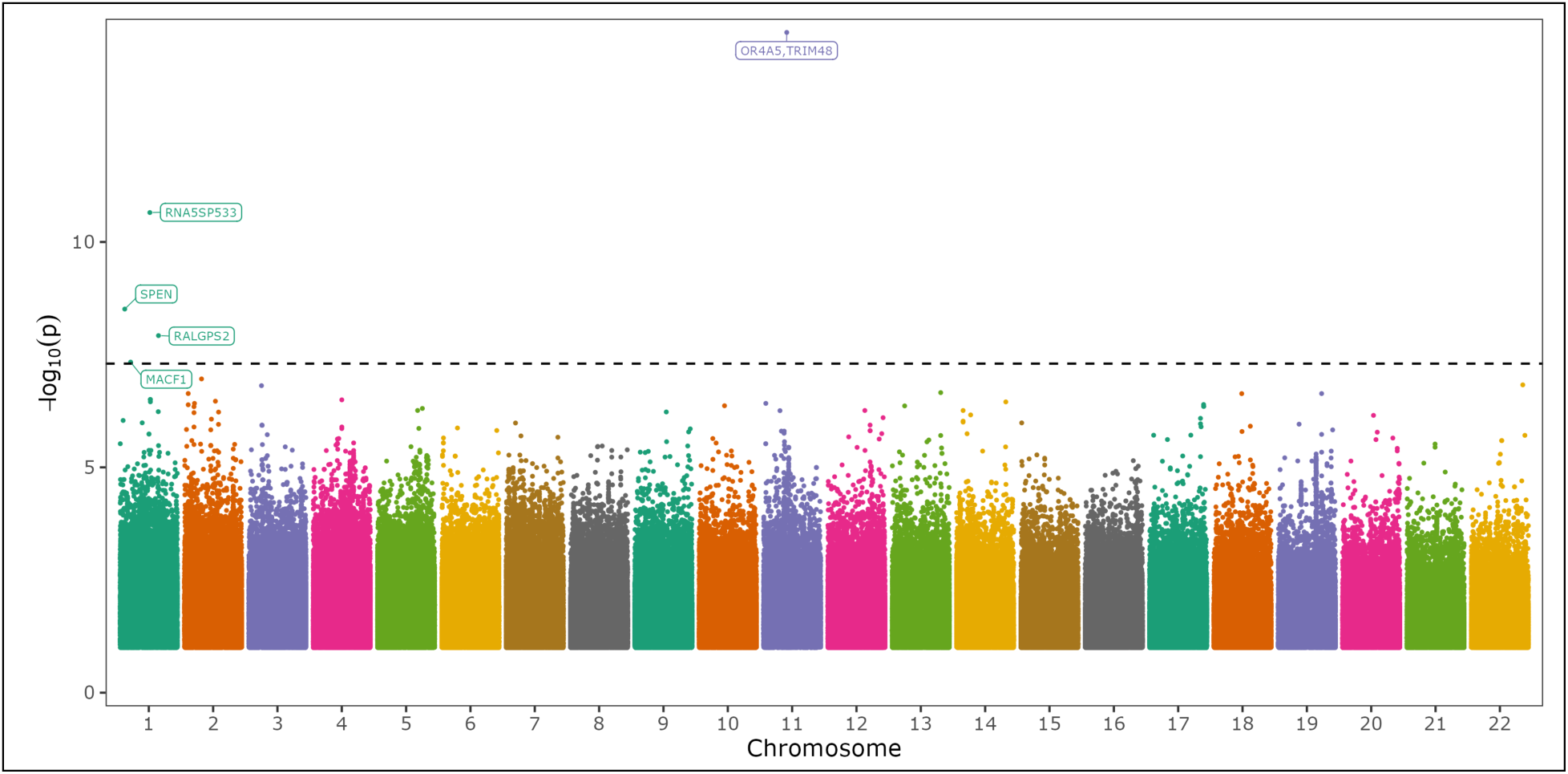
Manhattan plots of NIAGADS self-identified Hispanic participants and AoU studies conducted on participants who were matched to these NIAGADS participants using projected PCs, with or without age and sex covariates. The ZFYVE1 locus in (A) corresponds to rare variants proximal to PSEN1.

**Table 1.**
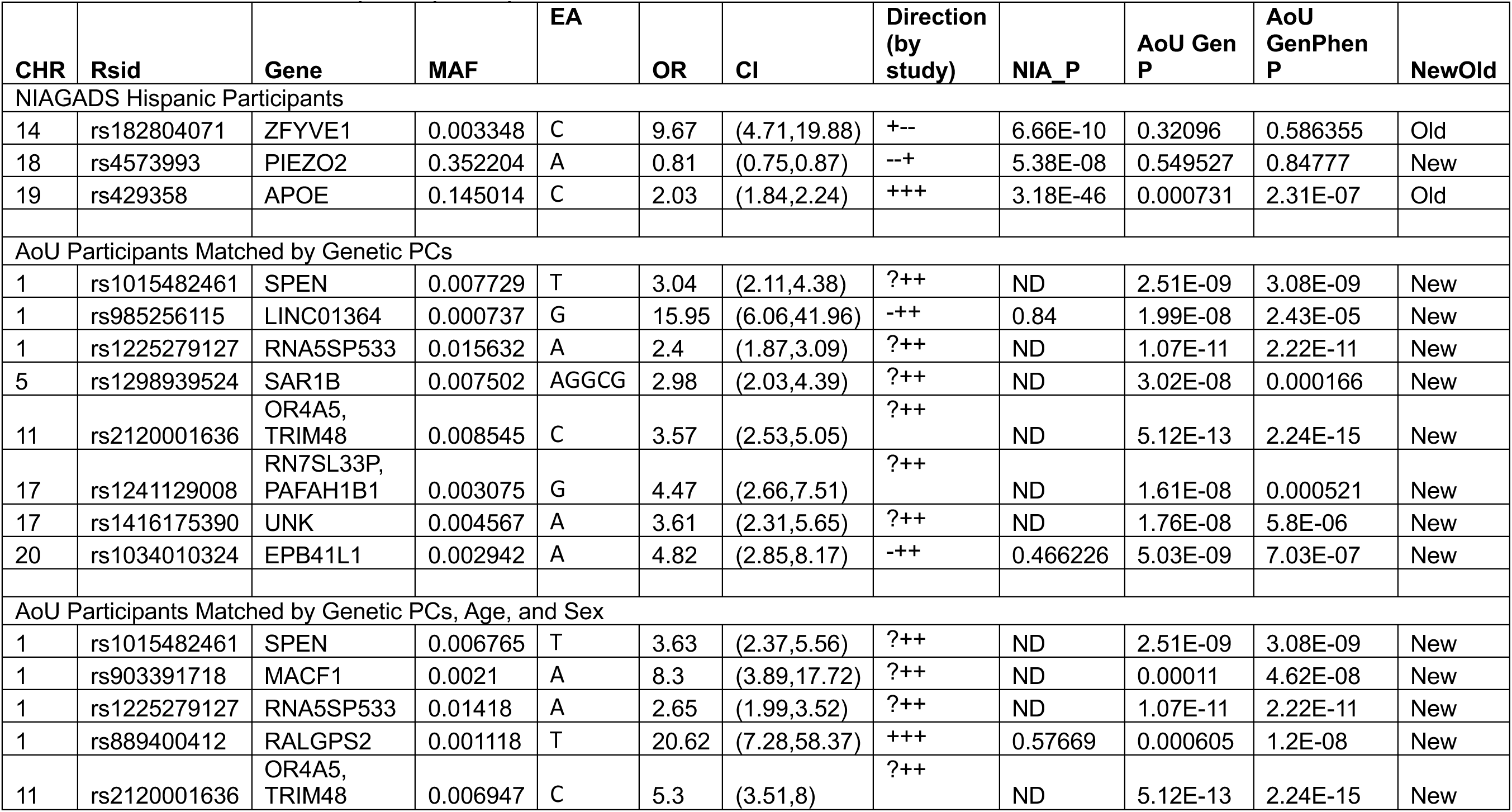
Lead variant statistics for each individual GWAS. AoU Gen P contains p-values for the AoU GWAS produced by matching AoU participants to NIAGADS participants who self-identified as Hispanic using solely the projected genetic PCs, with AoU GenPhen P produced by matching with PCs, age, and sex. While none of the AoU lead variants were nominal in the NIAGADS Hispanic participant GWAS, several of the loci contained non-lead variants that were.

### All of Us Participants Matched to NIAGADS GWAS

We next performed a GWAS in a genetically matched cohort from AoU, where genetic matching was performed with and without age and sex covariates. Lambda values for genomic inflation for both AoU GWAS were below 1.0 (**Supplemental Figure 1**). The *APOE* locus yielded a p-value of 2.3 * 10^-7^ in AoU when including age and sex in the matching schema, and a less significant p-value (p=0.0007) when matching solely on genetic PCs. (**Table 1**). For AoU participants matched with solely PCs, the GWAS yielded 8 novel loci (1 common, 7 rare), which were proximal to the gene encoding the (*SPEN)*, long intergenic non-protein coding RNA 1364 (*LINC01364*), 5S Ribosomal Pseudogene 533 (*RNA5SP533*), Secretion Associated Ras Related GTPase 1B (*SAR1B*), Olfactory Receptor Family 4 Subfamily A Member 5/ Tripartite Motif Containing 48 (*OR4A5*/*TRIM48*), RNA, 7SL, cytoplasmic 33, pseudogene/ Platelet Activating Factor Acetylhydrolase 1b Regulatory Subunit 1 (*RN7SL33P*/*PAFAH1B1*), Unk Zinc Finger (*UNK*), and Erythrocyte Membrane Protein Band 4.1 Like 1) (*EPB41L1*) (**Figure 3**, **Table 1**). The genome-wide significant lead variants from AoU were either not detected or not significant (**Table 1**). For AoU participants matched by PCs, age, and sex, a smaller cohort, we identified five new loci (1 common, 4 rare), of which two were also genome-wide significant in the AoU cohort matched by purely PCs. These loci were proximal to *SPEN*, Microtubule Actin Crosslinking Factor 1 (*MACF1*), *RNA5SP533*, Ral GEF with PH domain and SH3 binding motif 2 (*RALGPS2*), and *OR4A5*/*TRIM48* (**Figure 3**, **Table 1**). While there was extensive overlap of subjects in the two AoU sub-cohorts, each lead variant was nominally significant in the other (**Table 1**).

Focusing on the genes that were genome-wide significant in both AoU sub-cohorts, *SPEN* contains a lead, rare variant predicted to increase AD risk in both AoU sub-cohorts. *SPEN* is enriched in brain and plays a role in regulating gene transcription, believed mediated by hormonal signaling^15^. *SPEN* has not been previously linked to dementia, but haploinsufficiency in the gene has been linked to intellectual disability and brain anomalies, particularly in females^18^. *RNA5SP533*, a gene 230 kb from the lead variant in a common variant predicted to increase AD risk, is a ribosomal pseudogene^19^, with variants previously linked to breast cancer and anxiety^20^. Copy-number variants proximal to the olfactory gene, *OR4A5*^15^, have been associated with amyotrophic lateral sclerosis^21^. Finally, *TRIM48* plays a role in cell death related to oxidative stress, and a prior study has linked the gene to AD when studying individuals in extreme quantiles of an AD PRS^22^, a method that some believe increases the risk of false positives^23^.

### Meta-analyses of NIAGADS Sub-cohort and Matched AoU Participants

We next carried out a meta-analysis of the results from our NIAGADS sub-cohort with each matched AoU subpopulation. Genomic inflation remained controlled, with inflation factors less than 1 (**Supplemental Figure 1**). To focus on generalizable results, we focused on variants observed in both studies. This excluded the genome-significant loci we observed in the AoU GWAS, while yielding additional novel loci in each AoU sub-cohort that were also nominally significant in our NIAGADS sub-cohort. For a rare variant with a MAF of 0.001, 0.003 and 0.005 and a combined sample size of 53,260 (n_cases_=12,038, AoU matched by genetic PCs), we had at least 80% power to identify variants with a genotype relative risk (GRR) of 2.64, 1.81 and 1.59, respectively. If we used a combined sample size of 33,969 (n_cases_=7,192, AoU matched by genetic PCs, age and sex), the range of GRRs was 3.36, 2.11 and 1.81.

In both meta-analyses, the *APOE* locus was reproduced with supporting statistics across studies (**Figure 4**, **Table 2**). In addition, a rare variant in Regulator Of G Protein Signaling 6 (*RGS6)*, appeared in both meta-analyses, however it was in LD and < 1 Mb from PSEN1 mutations. In both meta-analyses, rs374043832 near *RGS6*, had a p-value just beyond nominal significance (p = 0.051 and p = 0.051) in the AoU analyses (**Table 2**). We also identified two novel variants in the meta-analysis employing AoU participants matched by genetic PCs, all rare, proximal to UBX Domain Containing Tether For SLC2A4 (*ASPSCR1*), and Erythrocyte Membrane Protein Band 4.1 Like 1 (*EPB41L1*) (**Table 2**), with the locus proximal to *ASPSCR1* being nominally significant in both NIAGADS and AoU (**Table 2**). This variant was present in 24/9479 cases (with one affected carrier being homozygous) and 193/35314 controls in AoU (matched by genetic PCs) and 23/2559 cases and 23/5908 controls in our NIAGADS cohort. We also observed one rare novel loci in the meta-analysis employing AoU participants matched by genetic PCs, age, and sex, proximal to Small nucleolar RNA SNORA40/Ganglioside Induced Differentiation Associated Protein 2 (*RF00561*/*GDAP2*) (**Table 2**). The locus proximal to *GDAP2* was nominally significant in NIAGADS and the AoU sub-cohort (**Table 2**). This variant was present in <20/4633 cases and 37/20869 controls in AoU (matched by genetic PCs, age, and sex) and 15/2559 cases and 15/5908 controls in NIAGADS. These loci, which were nominally significant, or nearly nominally significant, in both studies will be referred to as “higher confidence loci” (all loci except *EPB41L1*).

**Figure 4.**
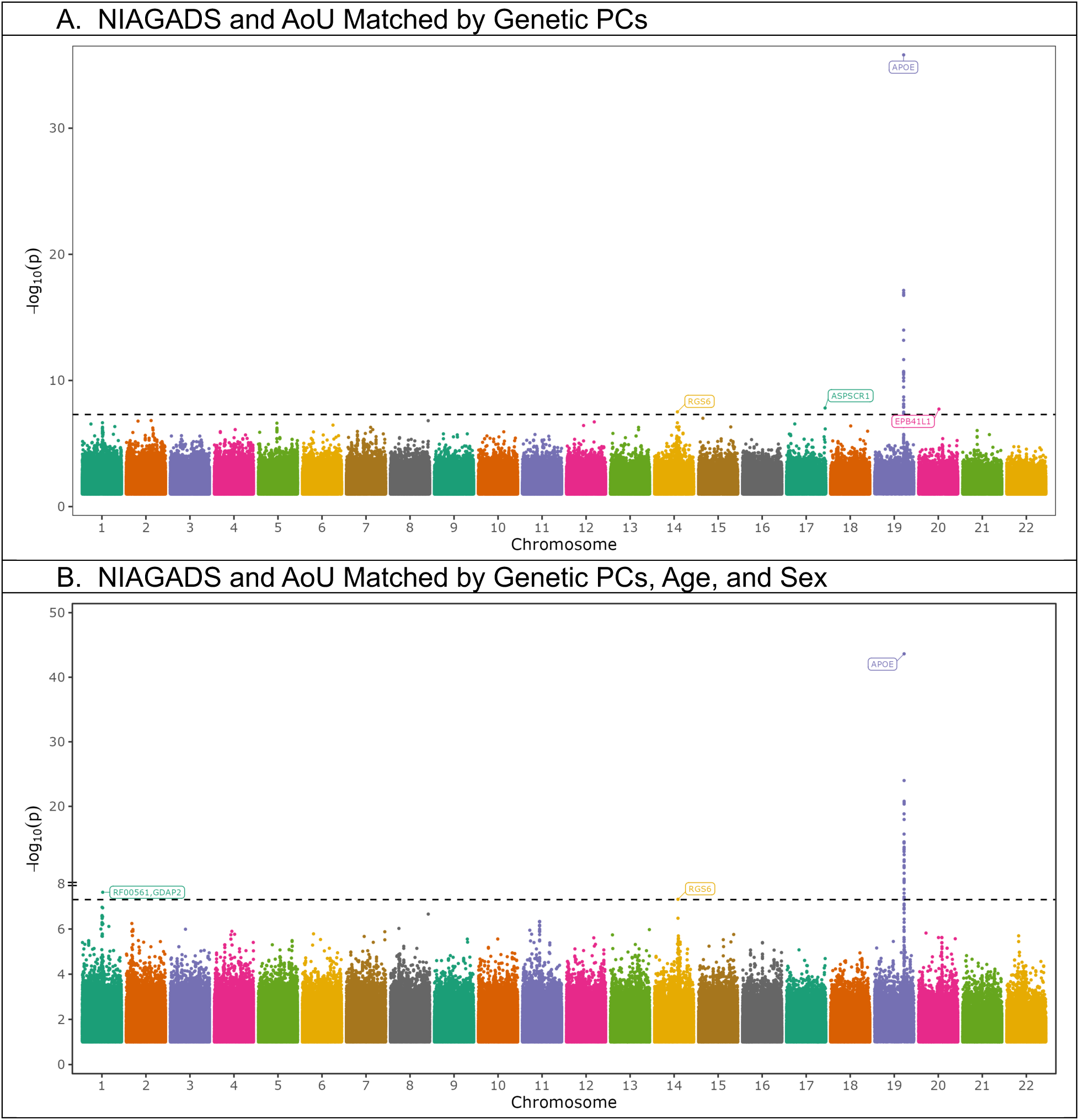
Manhattan plots for meta-analyses of NIAGADS participants who self-identified as Hispanic and matched All of Us cohorts, filtered for variants that occurred in both datasets.

**Table 2.** Lead variant statistics for each meta-analysis. Gen represents AoU participants matched to NIAGADS participants who self-identified as Hispanic using projected principal components. GenPhen represents AoU participants matched by these projected genetic PCs, age, and sex. Direction solely refers to the NIAGADS, AoU matched by genetic principal components, and AoU matched by genetic principal components and phenotypic traits.

| CH<br>R | Rsid | Gene | MAF | E<br>A | O<br>R | CI | Directio<br>n (by<br>study) | Meta<br>Gen<br>P | Meta<br>GenPhe<br>n P | NIA P | AoU<br>Gen P | AoU<br>GenPhe<br>n P | NewOl<br>d |
| --- | --- | --- | --- | --- | --- | --- | --- | --- | --- | --- | --- | --- | --- |
| NIAGADS HISP and AoU Participants Matched by Genetic PCs |  |  |  |  |  |  |  |  |  |  |  |  |  |
| 14 | rs374043832 | RGS6/PSEN<br>1 | 0.00<br>2 | A | 5.4 | (3.0,9.8) | +++ | 3.08E<br>-08 | 4.84E-08 | 4.86E-08 | 0.050 | 0.09 | Old |
| 17 | rs192423465 | ASPSCR1 | 0.00<br>3 | T | 3.3 | (2.2,5.0) | +++ | 1.52E<br>-08 | 4.2E-05 | 8.3E-3 | 1.95E-07 | 0.001 | New |
| 19 | rs429358 | APOE | 0.13<br>7 | C | 1.6 | (1.5,1.7) | +++ | 1.54E<br>-36 | 2.36E-44 | 3.2E-46 | 0.00073<br>1 | 2.31E-07 | Old |
| 20 | rs103401032<br>4 | EPB41L1 | 0.00<br>3 | A | 4.4 | (2.6,7.5) | -++ | 1.85E<br>-08 | 2.39E-06 | 0.47 | 5.03E-09 | 7.03E-07 | New |
| NIAGADS HISP and AoU Participants Matched by Genetic PCs, Age, and Sex |  |  |  |  |  |  |  |  |  |  |  |  |  |
| 1 | rs935208076 | RF00561,<br>GDAP2 | 0.00<br>1 | C | 8.1 | (3.9,17.0<br>) | +++ | 1.98E<br>-06 | 2.37E-08 | 0.00088<br>2 | 0.00067<br>4 | 1.82E-06 | New |
| 14 | rs374043832 | RGS6/PSEN<br>1 | 0.00<br>2 | A | 5.5 | (3.0,10.2<br>) | +++ | 3.08E<br>-08 | 4.84E-08 | 4.86E-08 | 0.051 | 0.08825<br>5 | Old |
| 19 | rs429358 | APOE | 0.13<br>7 | C | 1.7 | (1.6,1.8) | +++ | 1.54E<br>-36 | 2.36E-44 | 3.18E-46 | 0.00073<br>1 | 2.31E-07 | Old |

*ASPSCR1*, which contains an intronic variant in the mutually nominally significant locus, is enriched among pathways with roles in brain immune response and modulates glucose transport by interactions with Glucose Transporter Type 4 (GLUT4)^15^. This gene has been previously observed to be significantly less expressed in individuals with AD from data contained in the Gene Expression Omnibus database^24^. *GDAP2*, which is roughly 50 kb from a rare locus that was mutually nominally significant, is enriched among gene pathways with roles in transcription, previously implicated in spinocerebellar ataxia 27^15^, which includes dementia-like phenotypes^25^. *RGS6*, which contains an intronic variant in the nearly mutually nominally significant locus, is enriched among mixed processes in neurons, including neuronal signaling in single cells^15^. It displays enriched expression in excitatory and inhibitory neurons as well as astrocytes with roles involving inhibition of signal transduction through GTPase pathways^15^. RGS6 has been previously linked Parkinson disease^26^.

### Reproduction of Loci that were Nominally Significant in Both Datasets in Broader Cohorts

Having matched AoU participants by genetic and phenotypic similarity to our NIAGADS sub-cohort, we next set out to determine whether the results could be generalized to broader populations. We evaluated our higher confidence loci in our prior AoU study including all participants^27^, all AoU participants with admixed American genetic ancestry, AoU participants not in the matched cohorts, and in NIAGADS when studying all participants (**Supplemental Table 2**). We also attempted validation of our genome-wide significant variants in the Massachusetts General Brigham biobank, UK Biobank, and the newest release of NIAGADS after excluding participants we used in this study. We studied all these NIAGADS participants, also studying subcohorts, including new participants who identified as Hispanic and those with African, admixed American, and European genetic ancestry (**Supplemental Table 2**). All variants were nominally significant in the all-participant AoU GWAS, with only rs374043832 in *RGS6* being nominally significant in the all-participant NIAGADS study (**Supplemental Table 3**). When we evaluated variants for association with AD-by-proxy in AoU participants, who were not matched to the NIAGADS sub-cohort, only rs192423465 in *ASPSCR1*, was reproduced, but limited to AoU participants who did not meet the threshold for being matched to the NIAGADS sub-cohort when employing genetic PCs, age, and sex (**Supplemental Table 3**). There were 1,811 AD-by-proxy cases in AoU participants that did not meet the threshold for matching by solely genetic PCs and 7,044 cases for participants who did not meet matching criteria by genetic PCs, age, and sex. Aside from rs374043832 (*RGS6*) in AoU participants who were logged as having admixed American genetic ancestry, these variants were not significant or detected when we evaluated other populations, including AoU participants with Admixed American or African genetic ancestry, UK Biobank, or African American and Asian NIAGADS participants^27^. Several of our novel variants were validated in other independent cohorts, including rs4573993 in *PIEZO2* in MGB Biobank with matching effect direction (p = 0.006) and rs889400412 in *RALGPS2* in the updated NIAGADS cohort after excluding this study’s participants in the all participants cohort (p = 0.04) and European genetic ancestry cohort with matching effect direction (p = 0.006) (**Supplemental Table 3**). We found that several of the hits we identified that were already known were validated with rs374043832 in *RGS6/PSEN1* associated with AD in the new NIAGADS release’s Hispanic participants after excluding those in this study with matching effect direction (p = 0.001) and admixed American NIAGADS participants (p = 0.009) (**Supplemental Table 3**). rs192423465 in *ASPSCR1* in the updated NIAGADS Hispanic participant cohort after excluding this study’s participants had an opposite effect direction with nominal significance (p = 0.04) and rs182804071 in *ZFYVE1* in NIAGADS African ancestry participants had an opposite effect direction with nominal significance (p = 0.006) (**Supplemental Table 3**).

### Differential Gene Expression of Genes Proximal to Novel Loci

To further clarify the role of the genes proximal to our higher confidence loci, we used data from gene differential expression in brain single-cell populations for individuals with and without AD and pathological hallmarks of AD^28^. *ASPSCR1* was significantly less expressed in excitatory neurons when comparing individuals with pathological hallmarks of AD to those with and without cognitive impairment or comparing groups 3 and 4 (**Table 3**). *GDAP2* was significantly more expressed in excitatory neurons in individuals with pathological evidence of AD as compared to individuals without such evidence (**Table 3**).

**Table 3.** Differential gene expression for meta-analysis loci that were nominally significant in AoU and NIAGADS by presence of cognitive impairment or pathological evidence of disease. Exc represents excitatory neurons. Group comparisons (n v m) include group 1 (no AD pathological evidence or cognitive impairment), group 2 (no pathological evidence with cognitive impairment), group 3 (pathological evidence without cognitive impairment), group 4 (pathological evidence and cognitive impairment). Scores (xN) represent the number of unique cell subpopulations enriched for the cognate gene. Plus and minus signs correspond to whether the log-fold change was positive or negative for the comparison.

| CHR | Rsid | Gene | MAF | Cell |
| --- | --- | --- | --- | --- |
| 17 | rs192423465 | ASPSCR1 | 0.0026 | (ASPSCR1) Exc 4v1 (x4) ----<br>(ASPSCR1) Exc 4v3 (x1) - |
| 1 | rs935208076 | RF00561,<br>GDAP2 | 0.0011 | (GDAP2) Exc 3v1 (x2) ++ (GDAP2)<br>Exc 4v1 (x1) + |

We next evaluated these associations in additional analyses available from Mathys et al. 2023, which evaluated differential gene expression by severity of cognitive impairment, accounting for the extent of AD-characteristic pathological hallmark. In these studies, more excitatory neuron subpopulations demonstrated differential gene expression according to pathological features (**Table 4**)^28^. While *ASPSCR1* was significantly less expressed in individuals with no to mild to severe cognitive impairment in the setting of general AD pathology (group 1 vs 2 vs 3), eight subpopulations of excitatory neurons were downregulated in participants with mild cognitive impairment versus none when considering one’s amyloid burden and six subpopulations when considering neuritic plaque burden (**Table 4**). *GDAP2* was significantly upregulated across diverse pathological hallmarks when comparing by degree of cognitive impairment (**Table 4**).

**Table 4.**
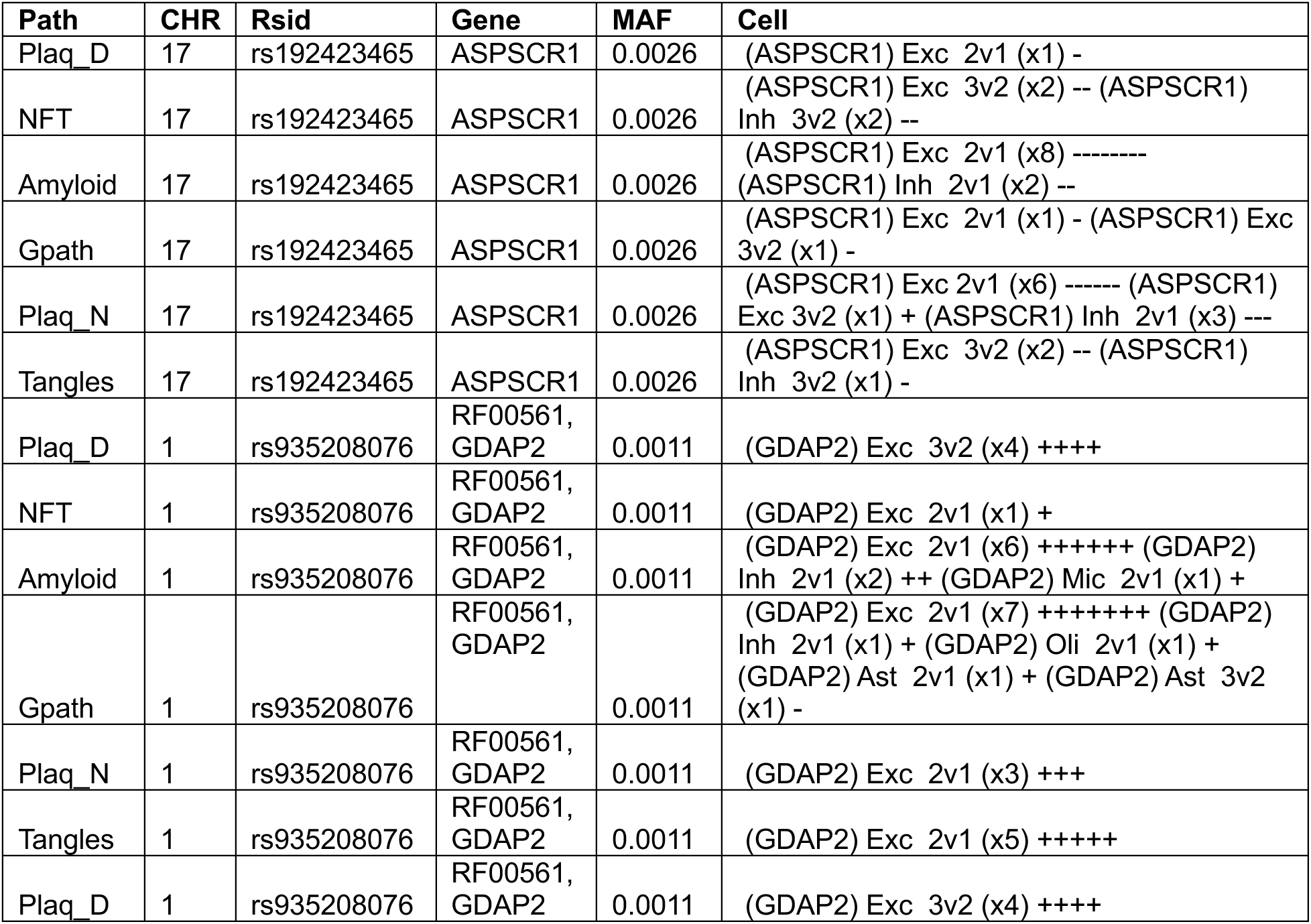
Differential gene expression for meta-analysis loci that were nominally significant in AoU and NIAGADS, by severity of cognitive impairment and specific pathological hallmarks of disease. For progression-relevant outcomes, amyloid represents amyloid burden, Gpath global AD pathology, NFT neurofibrillary tangle burden, Plaq D diffuse plaque burden, Plaq N neuritic plaque burden, tangles tangle density. For cells, Exc represents excitatory neurons, Inh inhibitory neurons, Oli oligodendrocytes, Ast astrocytes, 2v1 represents comparison of gene expression between individuals with mild cognitive impairment to those with no cognitive impairment, 3v2 comparing individuals with AD dementia and mild cognitive impairment. Scores represent the number of unique cell subpopulations enriched for the cognate gene with signs representing the direction of change in expression in each subpopulation.

## Discussion

In this study, we demonstrated how one can identify novel disease loci by matching subjects from smaller disease-focused cohorts to large biobanks while controlling for bias and population stratification, even for populations that are historically diverse. Projecting genetic data and matching participants by genetic PCs, we reproduced known AD loci and discovered new loci. The *APOE* SNP rs429358 was genome-wide significant in NIAGADS, however in AoU it showed less significant p-values, when subjects were matched by genetic PCs, age, and sex (p = 2.3 * 10^-7^), or by genetic PCs only (p = 7.3 * 10^-4^). This could be attributed to the potential noisiness of the less than optimal AD-by-proxy phenotype, especially for younger subjects^27^.

When age and sex were added to the matching covariates, older AoU subjects were selected and the *APOE* signal became more pronounced. In NIAGADS, we also identified a novel association of AD with a variant in *PIEZO2*, which was protective with a p-value just beyond genome-wide significance (p = 5.4 * 10^-8^). *PIEZO2* is a homolog of *PIEZO1*, which has previously been shown to influence clearance of Aβ^15^.

Meta-analyses yielded two novel rare loci, with lead variants rs192423465 (*ASPSCR1*), and rs935208076 (*GDAP2*) that were nominally significant in both cohorts and the broader AoU population (**Table 2**). rs192423465 was also reproduced in AoU participants who failed to meet inclusionary criteria when matching by genetic PCs, age, and sex (**Supplemental Table 3**). The genomic inflation factor for these meta-analyses were less than 1.

*ASPSCR1* and *GDAP2* have been linked to other neurodegenerative disorders with pathogenic mechanisms relevant to AD. *ASPSCR1* has been previously linked to amyotrophic lateral sclerosis and inclusion body myopathy with early-onset Paget disease and frontotemporal dementia^29^. ASPSCR1 plays a role in intracellular protein transport and, in particular, glucose homeostasis and transport. ASPSCR1 is a chaperone protein that serves to mitigate protein aggregation and misfolding. Future studies are needed to assess whether ASPSCR1 affects aggregation of hyper-phosphorylated tau and amyloid beta^30^. *GDAP2* mutations have been observed in individuals with autosomal recessive cerebellar ataxia, specifically spinocerebellar ataxia type 27, which may present with progressive spasticity and dementia^25^. This gene normally plays a role in cellular response to stress, lysosomal function, retinoic acid response, cerebral cortical atrophy, hippocampal excitatory synaptic activity, and has been linked to gliosis, a key feature of AD^19,31,32^ . Finally, RGS6 (near *PSEN1*), acts as a GTPase-activating protein and plays roles RGS6 has been previously associated with Parkinson disease^26^.

With regard to limitations of this study, we note that AD-by-proxy is used to determine affection status in the AoU biobank, with limited subjects diagnosed by clinical AD. Although the AD-by-proxy definition can potentially introduce biases^33^, it has been strongly correlated with AD status and increases statistical power, especially in population-based biobanks^34^. Its results, particularly in AoU, must be carefully curated and evaluated, as we demonstrated in a prior manuscript^27^. Here, we aimed to mitigate possible bias by initially focusing on signals that were nominally significant with clinical AD from NIAGADS. While we considered participants with first-degree relatives with AD or grandparents with AD, the latter being 25% genetically similar to the participant versus the 50% of first-degree relatives, we used this definition given precedents set by other studies^35,36^. Future validation studies of identified signals in other large Latino cohorts with clinical AD are warranted. We also note that we did not analyze sex chromosomes. The locus at *RGS6* is proximal to *PSEN1*, being less than 1 Mb away from rs63750082 (p.Gly206Ala). The rare variants shared most of the carriers in NIAGADS although showing modest LD (D’ = 0.1). While we emphasized variants that were genome-wide significant and nominal in both of our datasets while implementing quality control to focus on variants present in an adequate number of participants, with power calculations suggesting adequate power, they were rare variants with results that could be susceptible to bias. Some of these results, both from GWAS and meta-analysis, were also not validated on additional independent cohorts, which underscores the role of cohort on generalization of results.

## Conclusion

In meta-analyses matching subjects from smaller disease-focused cohorts to large biobanks, while controlling for bias and population stratification, we identified two novel loci associated with AD, *ASPSCR1* and *GDAP2*, all driven by rare variants. In addition, in NIAGADS subjects who self-identified as Hispanic, we identified a common variant in (*PIEZO2*), that was protective for AD with a p-value just beyond genome-wide significance (p = 5.4 * 10^-8^). Importantly, we show that population stratification in heterogeneous populations can be mitigated in meta-analyses by matching participants using projected principal components, enabling the use of large, multiethnic, and, importantly, diverse biobank cohorts in combination with smaller disease-specific cohorts.

## Methods

### Cohorts

The NIAGADS dataset includes harmonized phenotypes and sequencing data from cohorts sequenced by the Alzheimer’s Disease Sequencing Project and other AD and Related Dementia’s studies. Full details can be found on NIAGADS (https://dss.niagads.org/datasets/ng00067/) and elsewhere^37^. The NG00067.v9 release was used for this paper, which included 26,243 participants with an AD diagnosis. In addition, we used NG0067.v14 for validation in subjects, which were not included in the v9 release. The All of Us (AoU) cohort studies traditionally underrepresented individuals in biomedical research from the United States of America, offering 245,388 individuals’ short-read whole genome sequencing calls in release 7 with information from surveys, wearables, physical measurements taken at the time of participant enrollment, and electronic health records (EHRs)^38,39^. The Mass General Brigham (MGB) Biobank (https://biobank.massgeneralbrigham.org/) is a hospital-based research cohort comprising over 135,000 individuals recruited from multiple institutions within the MGB healthcare system, the parent organization of Massachusetts General Hospital and Brigham and Women’s Hospital^40^. All participants in this biobank provided informed consent for biospecimen collection, access to electronic health records (EHRs), and genomic analysis. The cohort includes individuals of diverse ancestries and clinical backgrounds, enabling broad applicability to translational research. Whole Genome Sequencing (WGS) was performed for a subset of participants, including all ICD-10 Alzheimer’s disease cases and older control subjects which included a substantial proportion of APOE carriers. WGS was run at the MGB PM Biobank Genomics Core using Illumina TruSeq DNA PCR-Free library construction kit with sequencing using NovaSeq 6000. Variant calling was performed using Illumina’s DRAGEN pipeline. The UK Biobank, a long-term study based in the United Kingdom, has gathered an extensive array of healthcare data from 502,357 individuals. This includes short-read whole-genome sequencing data from 200,004 of these participants, which were employed in this study.

### Outcomes

In NIAGADS, cases were individuals meeting the NINCDS-ADRDA criteria for definite, probable, or possible AD, had documented age at death (for pathologically verified cases) or age at onset, and ApoE genotyping. Controls were individuals over age 60 and free of dementia. AoU and UKB cases are defined by individuals having an ICD-9 (331.0) or ICD-10 (G30.9) code for AD or history of dementia in a first-degree relative or grandparent. While prior studies have focused on individuals with affected parents for a proxy case^3^, with quantitative definitions reportedly limiting bias^41^, genetic similarity between any first-degree relative is comparable. Affected grandparents are minimum 25% genetically similar, with precedent for their inclusion^35,36^. A quantitative definition was not possible in AoU, contrary to in UKB, given that parental age and age of parental death were unavailable^41^.

### Genetic data quality control

For NIAGADS, WGS variant calls for biallelic variants in 34,438 individuals were downloaded for the NIAGADS dataset from NG00067.v9 release. We excluded technical replicates, keeping the sample with fewer missing variant calls, those with a missing AD diagnosis, outliers by HET/HOM ratio (6 standard deviations from the mean), subjects with greater than 5% sample missingness, subjects from pairs that were second degree relatives or closer per KING^42^ calculations. The final dataset had 26,243 subjects. We focused on individuals who self-identified as Hispanic (n = 8,467), excluding subjects who were outliers based on genetic PCs, having analyzed and meta-analyzed data for the full cohort in a prior paper^27^. In addition, we have used the “bigsnpr” package and followed their vignette (https://privefl.github.io/bigsnpr/articles/ancestry.html#ancestry-grouping) to cluster subjects into groups of predicted genetic ancestry (**Supplemental Table 1**). Briefly, we used a set of reference allele frequencies and corresponding PC loadings provided by this paper^43^. Next, we used Euclidean distance on the PC space to assign a subject to the closest one of the 21 reference groups. We further combined the 21 groups into 5 continental-based groups (reported in Supplemental Table 1).

For replication in the independent subjects from NG00067.v14 release, which were not included in the v9 release, we followed the same protocol as above but excluded the original self-identified Hispanic subjects. This new-subject-only cohort consisted of 32,377 total individuals, with 3,018 being self-identified Hispanic. We also used the same PC-based grouping method above to categorize the new subjects into three ancestry groups: African (n = 5,748), American (n = 1,652), and European (n = 20,062). were processed similarly.

For AoU, data was processed similarly to Willett et al. 2025^27^, which studied AD-by-proxy in AoU and UKB, using processing similar to others^44,45^. Full Hail variant dataset (VDS) files for AoU were filtered for 550 AoU-flagged samples and AoU-flagged variants, leaving 1,085,790,733 variants and 244,845 samples. We removed variants with more than 10% missingness or occurring in fewer than 20 individuals, given data-reporting privacy practices^3,45^.

For the UKB data, WGS files were initially converted from their original VCF format to biallelic PGEN format using PLINK2 (with original multiallelic variants being split using bcftools) to make them compatible with subsequent Regenie analysis. The files were filtered for variants with more than 10% missingness, samples with more than 10% missingness, Hardy-Weinberg p-values less than 1 x 10^-15^, variants occurring in less than 3 individuals, and spanning deletions^3,45^.

For MGB, WGS data on 3,227 individuals were used. We utilized the PC-based clustering method above to group the participants into ancestry categories. We kept only those subjects of European ancestry and were left with 3,142 individuals.

### Matching of NIAGADS participants with All of Us participants

Using the BigSnpR package^46^, we imported the raw genetic data for common (MAF ≥ 1%) variants meeting HWE and call rate criteria from our NIAGADS sub-cohort and all AoU participants. We then projected the genetic data into a shared space using the “bed_projectPCA” function from the same package, using the NIAGADS sub-cohort as the reference population and outputting the first 20 projected principal components. We obtained the projected PCs for our model using the “predict()” function on the projected results’ output’s SVD of the reference data for NIAGADS and the online augmentation, decomposition, and Procrustes (OADP) projection results of the model for AoU results. We then produced two models using the R package, MatchIt^47^. We used the first twenty projected PCs alone with AD case status as covariates against cohort as the outcome for the first model (Cohort ∼ PCs + AD status). The second model was identical, also including age and sex as covariates. We used a subclassification as our method, computing 500 subclassification groups. Distance was computed using a generalized linear model and the estimand used NIAGADS as the reference group, thus each NIAGADS participant had a weight of 1 in the output model. To select for AoU participants most similar to our NIAGADS sub-cohort, we isolated AoU participants for each model with a weight matching or exceeding the NIAGADS reference weight of 1. Code for this analysis is available in our Code Availability section.

### Genome-wide association analysis

For NIAGADS we performed a logistic regression (with option “firth-fallback”) for case/control status as implemented in Plink2, studying individuals of varying sub-cohorts, which included those self-reporting as Hispanic for this study^48^. We included sex, age, sequencing center, sample set and 5 Jaccard principal components with standardized variance as covariates^49^.

For AoU and UKB, Regenie v3.2.8 was used to conduct GWAS^45^. Covariates for the AoU analysis included age of enrollment, sex, and the first twenty principal components (PCs) for members of each tested cohort, determined using Plink2^44,48^.

Sequencing center information in AoU was unavailable^38^. Covariates for the UKB analysis included age at enrollment into UKB, sex, the first twenty PCs, and sequencing center^50^. Outcomes were defined, as above. Step 1 was accomplished with array data for participants in each matched subcohort, following similar methodology to another study that worked on AoU, filtering for variants with a MAF ≥ 1%, minimum allele count of 100, and variant missingness ≤ 10%^44^. This data was then pruned for independent variants using 100 kb windows with a single-base step-size and 0.1 r^2^ threshold, with sex chromosomes removed.

Step 2 was accomplished using whole-genome sequencing files, processed as described above, with array-based step 1 predictions. We used Firth penalized regression for variants with p < 0.01 and a minimum minor allele count of 20 for AoU. Step 2 for UKB employed a Firth penalized regression to variants with a P-value less than 0.05. Reported AoU GWAS summary statistics were filtered for any variant where the MAF x N for the minor allele was less than 20, per the AoU privacy policy. Following GWAS, we determined the HWE MIDP value, using Plink2, for every variant with a p-value ≤ 1* 10^-5^, excluding variants with a MIDP value ≤ 1* 10^-15^, in a single genetic ancestral population, similar to our prior paper^27^. We used a standard p-value threshold of 5* 10^-8^ as our threshold for genome-wide significance, which accounts for multiple-testing across all independent genetic variants^51^.

While we did not include summary statistics or Manhattan plots for our analyses on AoU participants who self-identified as Hispanic, information on these cohorts is available in our Supplementary materials (**Supplemental Table 1**). As expected, genomic inflation was high in GWAS data focused on AoU participants who self-identified as Hispanic. Inflation in that cohort was greatly improved by including genetic ancestry as a covariate, which motivated this paper’s methods.

### Meta-analysis

Meta-analyzed results were processed using METAL^52^ (the most recent version, released on 25 March 2011) using default settings, aside from using inverse standard error values as weights and outputting the average allele frequency across studies. The final results were filtered for genetic variants with a frequency amplitude less than 0.4 (or difference between the maximum and minimum allele frequency between studies^3^), variants that were genome-wide significant, variants that were detected in both the AoU and NIAGADS studies (a degrees of freedom value of 1), and variants that had a HWE p-value larger than 1* 10^-15^ in all AoU ancestry-stratified populations (EUR, AFR, AMR, EAS, SAS, or MID). Following this, we used Plink2 to clump every significant variant by locus in our study using AoU genetic data, using a threshold R^2^ of 1%^48^. We used a standard p-value threshold of 5* 10^-8^ as our threshold for genome-wide significance^51^.

### Variant gene annotations and differential gene expression

We used FAVOR to obtain the genes proximal to each genetic variant^53^. For variants that passed inclusionary criteria, we then evaluated their cognate gene’s differential expression in single brain cells for individuals with and without pathological and clinical features of AD using data from Mathys et al. 2023 using an FDR cutoff of 1%^28^. Their analysis incorporated single-cell sequencing of 2.3 million prefrontal cortex cells from 427 individuals with varying degrees of AD pathology and cognitive impairment. These cells were grouped to form 12 major categories with 54 high-resolution cell types, including excitatory neuron subtypes, inhibitory neuron subtypes, oligodendrocytes, oligodendrocyte precursor cells, astrocyte subtypes, immune cell types, and vascular cell types^28^.

### Power calculations

We used the Genetic Association Study Power Calculator, available from the University of Michigan, to calculate power (https://csg.sph.umich.edu/abecasis/gas_power_calculator/). We use case and control counts from our study (**Supplementary Table 1**), a significance level of 5X 10^-8^, an additive disease model, a disease prevalence of 11.1%, and used variant-level information for disease allele frequency and genotype relative risk.

## Data Availability

All data produced in the present study are available upon reasonable request to the authors.

## Acknowledgements

This work was supported by Cure Alzheimer’s Fund, the JPB Foundation (RET), and NIGMS T32GM007748 (JW). The All of Us Research Program is supported by the National Institutes of Health, Office of the Director: Regional Medical Centers: 1 OT2 OD026549; 1 OT2 OD026554; 1 OT2 OD026557; 1 OT2 OD026556; 1 OT2 OD026550; 1 OT2 OD 026552; 1 OT2 OD026553; 1 OT2 OD026548; 1 OT2 OD026551; 1 OT2 OD026555; IAA #: AOD 16037; Federally Qualified Health Centers: HHSN 263201600085U; Data and Research Center: 5 U2C OD023196; Biobank: 1 U24 OD023121; The Participant Center: U24 OD023176; Participant Technology Systems Center: 1 U24 OD023163; Communications and Engagement: 3 OT2 OD023205; 3 OT2 OD023206; and Community Partners: 1 OT2 OD025277; 3 OT2 OD025315; 1 OT2 OD025337; 1 OT2 OD025276. In addition, the All of Us Research Program would not be possible without the partnership of its participants. Access to individual-level data from the All of Us research program was obtained through an MGB-signed a data use agreement with All of Us (www.researchallofus.org/register/). The content is solely the responsibility of the authors and does not necessarily represent the official views of the National Institutes of Health. We value the participants who volunteered for both the UKB and AoU. We appreciate the assistance of Cynthia Morton and James Gusella for providing feedback for this study. The computations in this paper were run in part on the FASRC Cannon cluster supported by the FAS Division of Science Research Computing Group at Harvard University. Please refer to the Supplementary Note for full acknowledgements. Samples, genomic data, and health information for the MGB cohort were obtained from the Mass General Brigham Biobank, a biorepository of consented patients samples at Mass General Brigham (parent organization of Massachusetts General Hospital and Brigham and Women’s Hospital). Whole genome library prep and sequencing of samples was performed at the Mass General Brigham Biobank Genomics Core with funding from Cure Alzheimer’s Fund.

## Author Contributions (CRediT Author Statement)

Julian Willett: conceptualization, methodology, software, validation, formal analysis, investigation, data curation, writing – original draft, writing – review and editing, visualization, supervision. Mohammad Waqas: formal analysis, validation, data curation, writing – review and editing. Serhiy Naumenko: data curation, writing – review and editing. Kristina Mullin: project administration, data curation, writing – review and editing. Julian Hecker: validation, writing – review and editing. Christoph Lange: validation, resources, writing – review and editing. Lars Bertram: validation, writing – review and editing. Winston Hide: validation, data curation, resources, writing – review and editing. Rudolph Tanzi: conceptualization, validation, resources, writing – review and editing, supervision, funding acquisition. Dmitry Prokopenko: conceptualization, methodology, validation, formal analysis, resources, data curation, writing – review and editing, supervision, project administration, funding acquisition.

## Data availability

NIAGADS data access is available through the DSS NIAGADS under accession number: NG00067. The NIAGADS dataset contains data in part obtained from the Alzheimer’s Disease Neuroimaging Initiative (ADNI) database (adni.loni.usc.edu). As such, the investigators within the ADNI contributed to the design and implementation of ADNI and/or provided data but did not participate in analysis or writing of this report.

UKB data access is available through application at https://ukbiobank.dnanexus.com/landing. Access to individual-level data from the All of Us research program was obtained through an MGB-signed data use agreement with

All of Us (www.researchallofus.org/register/). This research was conducted using the UKB resource (application number 81874). Full GWAS data for this manuscript, edited to comply with privacy requirements for AoU, is available by request.

## Code availability

Code is publicly available on Github: https://github.com/juliandwillett/NIA_AOU_ProjPCs_Study.

---

**Supplemental Figure 1.**
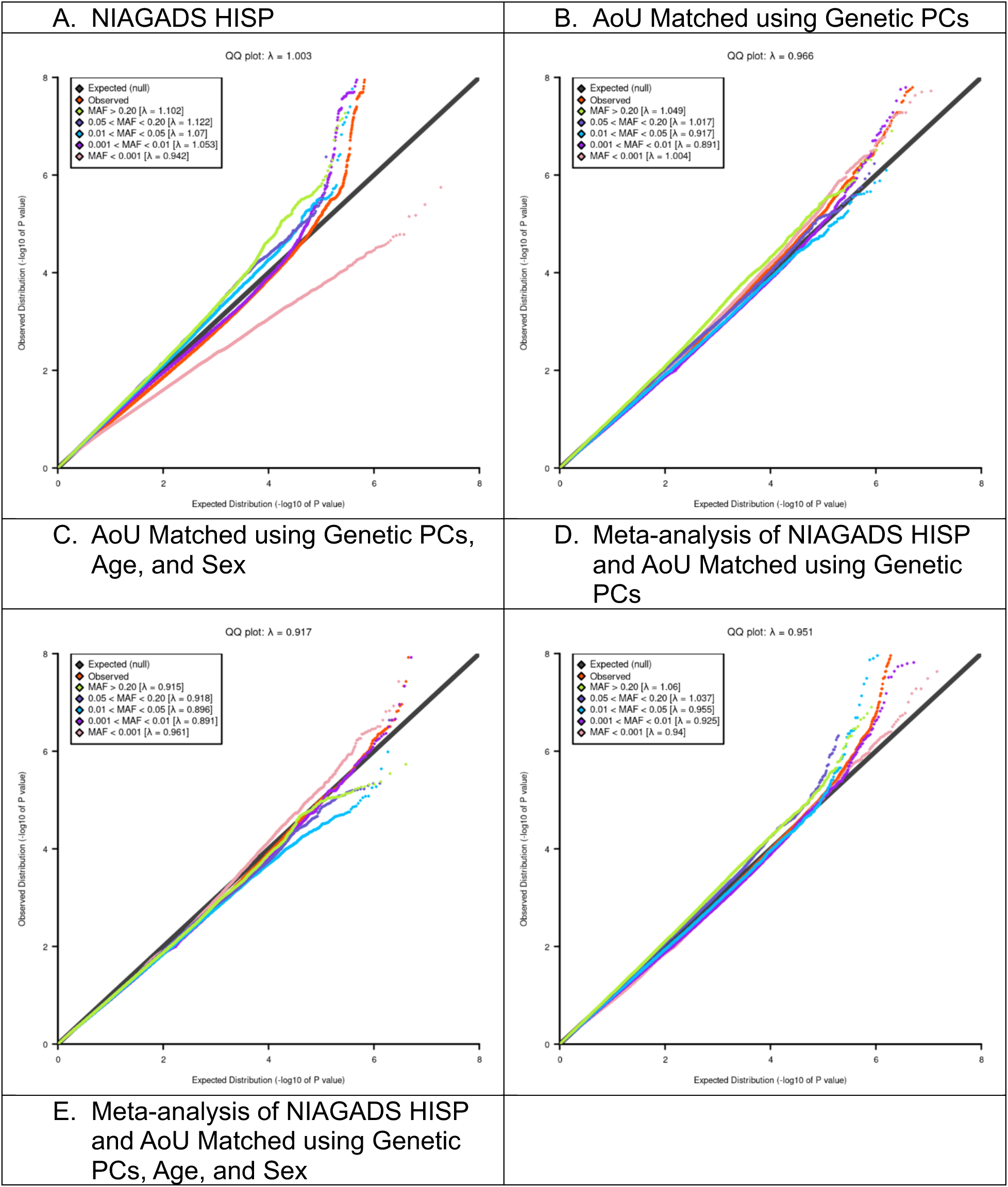
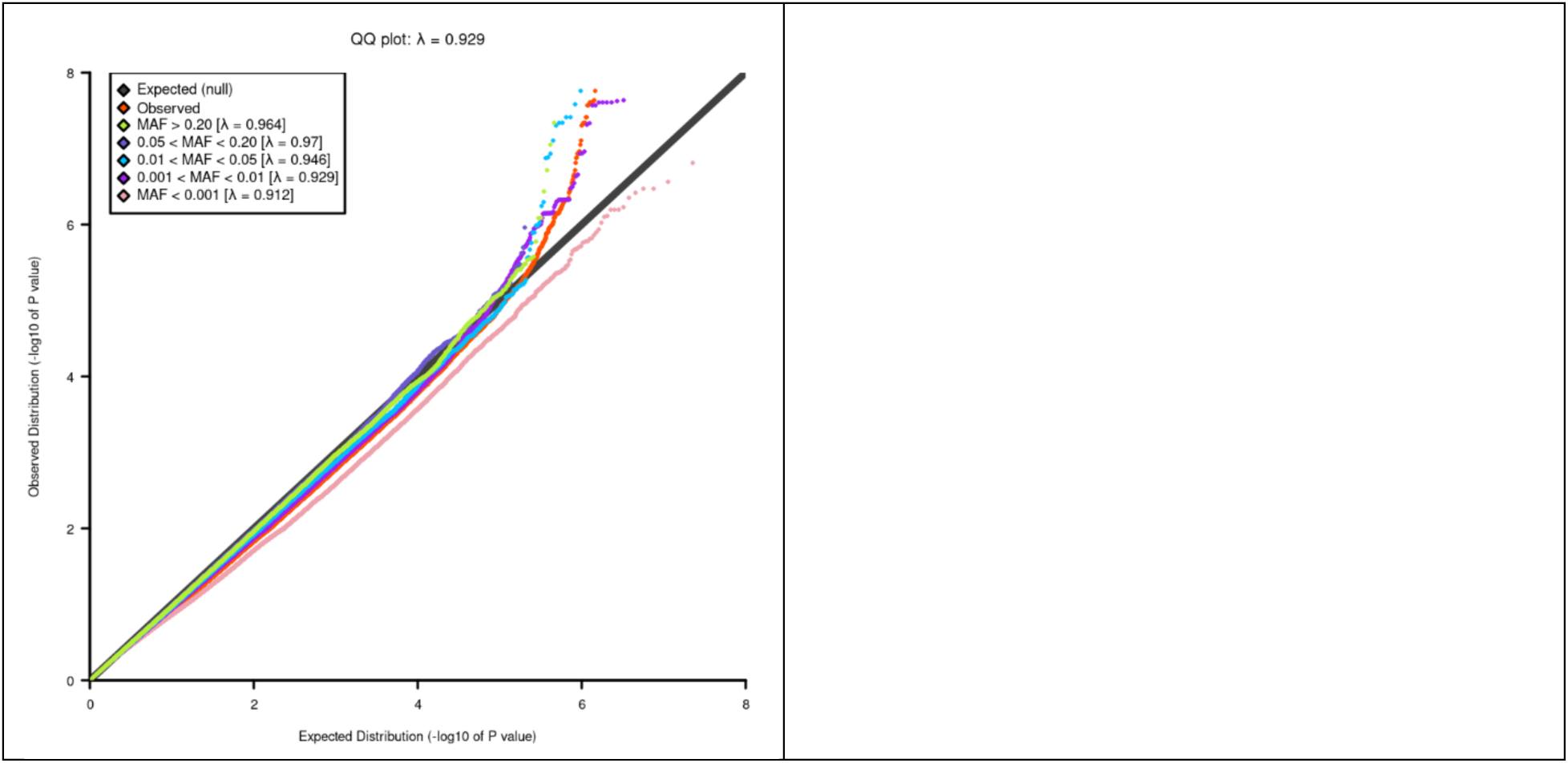
QQ plots, with genomic inflation factor, for each dataset.

**Supplemental Figure 2.**
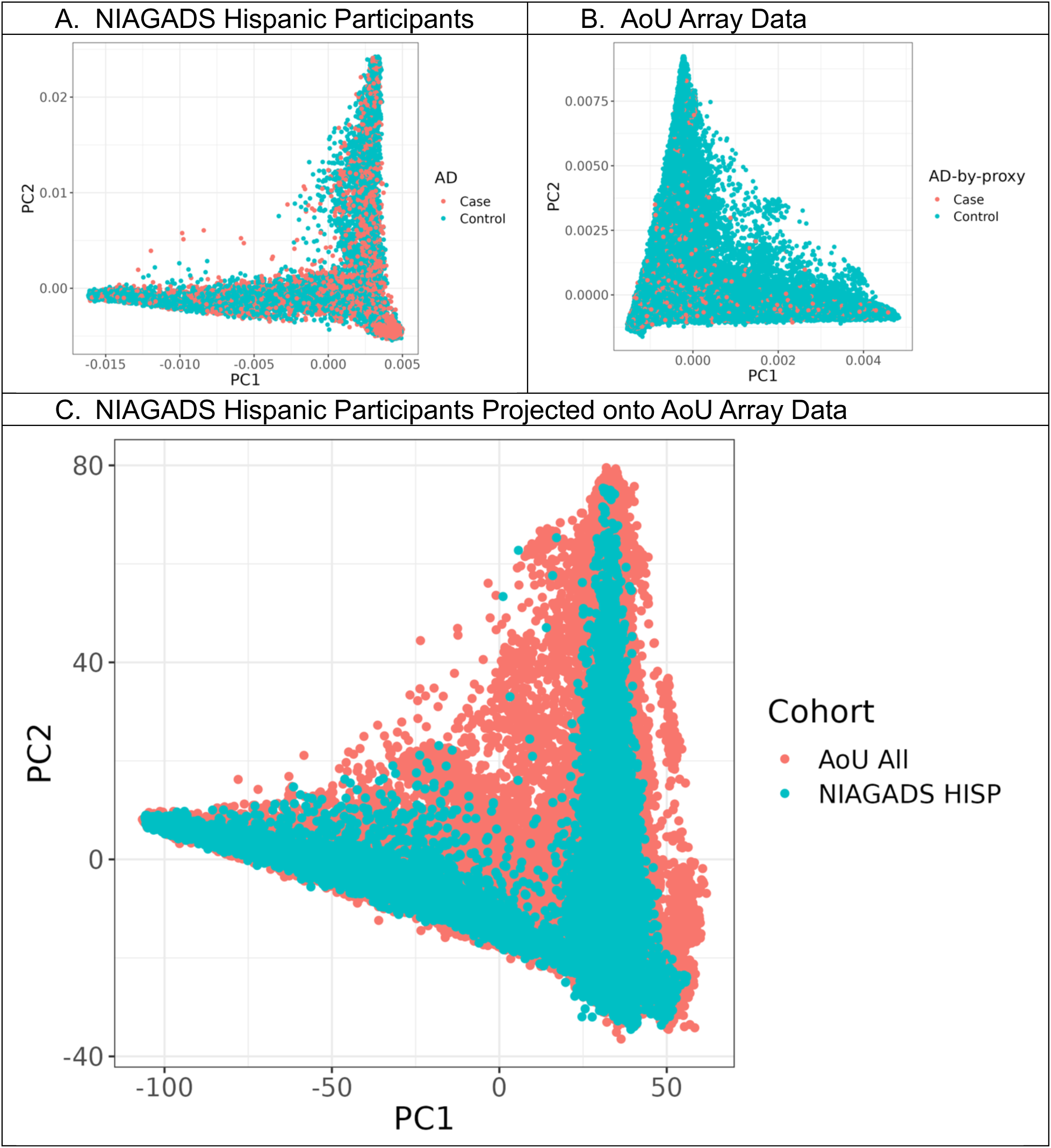
Principal component plots for each dataset.

**Supplemental Table 1.**
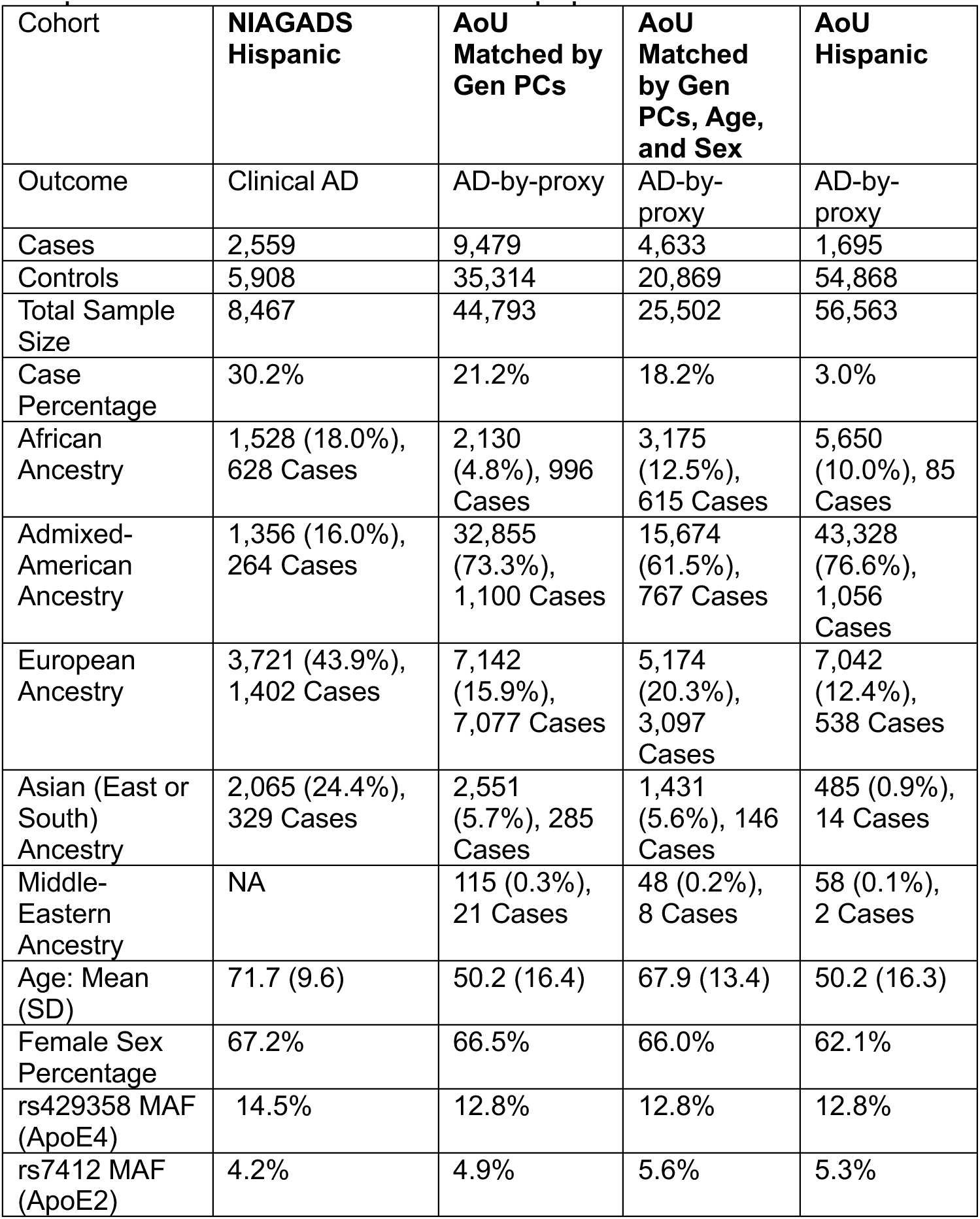
Demographic information for each cohort in our study. Single ancestral population percentages represent the proportion of the total sample size composed of the indicated ancestral population.

